# Randomized, double blind, placebo controlled, clinical trial to study co-administration of Ashwagandha on safety, immunogenicity, and protection with COVID-19 vaccine – A Study Protocol

**DOI:** 10.1101/2021.07.02.21259886

**Authors:** Arvind Chopra, Preeti Chavan-Gautam, Girish Tillu, Manjit Saluja, Swapnil Borse, Sanjeev Sarmukaddam, Susmita Chaudhuri, BCS Rao, Babita Yadav, Narayanam Srikanth, Bhushan Patwardhan

## Abstract

**Introduction:** The government of India has rolled out COVID-19 vaccine program for individuals who are 18 years of age and above and priority is being given to the elderly, and individuals with morbidity. Oxford-AstraZeneca COVID-19 vaccine (COVISHIELD™) is most widely used in India. A large number of Indian people have been consuming various traditional medicines in the hope of better protection against COVID-19 infection. Several studies have reported immunological benefits of Ashwagandha and its potential as vaccine adjuvant. We plan to study co-administration of Ashwagandha with COVISHIELD™ vaccine on safety, immunogenicity and protection.

**Methods and analysis:** We designed a prospective, randomized, double blind, parallel group, placebo controlled, two arm, exploratory study on healthy volunteers receiving the COVISHIELD™ vaccine. In addition to the two dose schedule of COVISHIELD vaccine as per national guidelines, participants will be administered 8gm Ashwagandha or placebo tablets respectively per day. Primary outcome measure is immunogenicity as measured by SARS-CoV-2 spike (S1) and RBD-specific IgG antibody titres. Secondary outcome measures are safety, protective immune response and quality of life measures. Adverse event following immunization will be monitored at each time throughout the study. Participants will be tracked on a daily basis with a user friendly mobile phone application. Following power calculation 600 participants will be recruited per arm to demonstrate superiority by a margin of 7% with 80% power. Study duration is 28 weeks with interim analysis at the end of 12 weeks.

**Ethics and dissemination:** Ethical approval was obtained through the Central and institutional Ethics Committees. Participant recruitment is expected to commence by August 2021. Results will be presented in conferences and published in preprint followed by peer-reviewed medical journals.

**Registration details:** Clinical Trial Registry – India (CTRI) Registration Number: CTRI/2021/06/034496. Date of Registration June 30, 2021.

**Strengths and limitations of this study:**

- Novel study to demonstrate effect of coadministration of immune adjuvant and COVID-19 vaccine on safety and immunogenicity.
- Randomised placebo controlled 28 weeks study with 80 percent power to demonstrate role of putative natural immunomodulator to augment the protection and reduce breakthrough infections
- State of art immune assays to measure specific antibodies to SARS-CoV-2 to demonstrate both persistent and late upsurge in immune response
- Daily tracking of participants using a study specific designed mobile app
- An interim analysis is planned to provide information on early immune response after first dose of vaccine
- Participants may be reluctant to donate blood repeatedly for immune assays; compliance with the test drug may be a challenge; asymptomatic infections may be missed; the study is not measuring cellular immune response.

## INTRODUCTION

The government of India has rolled out COVID-19 vaccine program for individuals who are 18 years of age and above and priority is being given to the elderly, and individuals with morbidity. The correlates of clinical protection against COVID-19 infection, and the immunological thresholds required for vaccine efficacy are not well established [1] [2]. Clinical studies have suggested a protective role for both humoral and cell-mediated immunity in recovery from SARS-CoV-2 infection. Antigen specific CD4+ T cells in COVID-19 patients are found to correlate with levels of antigen specific antibody responses [3]. ChAdOx1 nCoV-19 vaccination is reported to induce a predominantly Th1-type response [4].

The health-seeking pattern of the people suggests a large section of the population, particularly in India has been consuming various AYUSH preparations and botanical immunomodulators in the hope of better protection against COVID-19 infection. Registration of more than 1.3 crore health-seekers in the AYUSH Sanjivani App indicates this trend [5].

*Withania somnifera* (WS) also known as Ashwagandha in Ayurveda is a widely used medicinal plant. The classical Sanskrit language texts in Ayurveda describe Ashwagandha as Rasayana which means that it has therapeutic properties of being an immunomodulator, adaptogen and revitalizing agent [6] [7]. Various studies on Ashwagandha have reported its use in infection, inflammation and diseases such as arthritis and cancer [8]. In previous studies its selective-Th-1 upregulating activity was demonstrated in mouse models [9], which seems relevant in the management of COVID-19. Immunoadjuvant activity of Ashwagandha has been reported in an infection model when co-administered with DPT (diptheria, pertussis, tetanus) vaccine [10]. The long-term safety of Ashwagandha is well documented [11] [12] [13] [14] [15]. Based on available literature and empirical evidence, Ashwagandha is recommended by AYUSH ministry for management of asymptomatic and mild COVID-19 [16]. It is also being investigated in comparison with hydroxychloroquine in a randomized controlled multi-centric clinical study for its prophylactic activity against COVID-19 in high-risk health care workers. (CTRI/2020/08/027163, CTRI/2020/05/025332). Based on our earlier studies, we hypothesise that co-administration of Ashwagandha with COVID-19 vaccine may result in better safety, immunogenicity and protection.

We propose to study the effect of Ashwagandha co-administration with the COVID-19 vaccine (COVISHIELD™) on safety, immunogenicity and protection in healthy population. COVISHIELD™ consists of a replication-deficient chimpanzee adenoviral vector ChAdOx1, containing the SARS-CoV-2 structural surface glycoprotein antigen (spike protein; nCoV-19) gene [17] [18]. Considering that long-term immune response to the vaccine will have implications for prevention of reinfection, it is also of interest to study the effect of Ashwagandha on the sustenance of the vaccine response. This study will help to understand the potential of Ashwagandha to improve vaccine response on a long-term basis. The primary hypothesis is that a long-term administration of Ashwagandha along with COVISHIELD™ vaccine leads to a better protection against COVID-19 in healthy populations as compared to COVISHIELD™ vaccine alone and in terms of augmented immune-response, reduced incidence of breakthrough SARS-CoV-2 infections, good safety and tolerance. An important aim of the study is also to evaluate the salubrious effects of Ashwagandha on physical and mental health and quality of life.

## METHODS AND ANALYSIS

### Study design

This will be a prospective, randomized, double blind, parallel group, placebo controlled, multicentric study on healthy volunteers receiving the COVISHIELD™ vaccine with a superiority design. The trial will be conducted at multiple locations as shown in the CTRI. Consenting and eligible volunteers will be enrolled and randomized to either of the two study arms (Ashwagandha plus COVISHEILD™ vaccine or placebo plus COVISHIELD™ vaccine) as per the protocol. The participants will be jointly evaluated by an Ayurvedic and a modern medicine physician for several clinical and laboratory measures. An Ayurveda based clinical assessment will also be carried out. The various assessments to be carried out are shown in **Table 1** and **Table 2**.

**Table 1:**
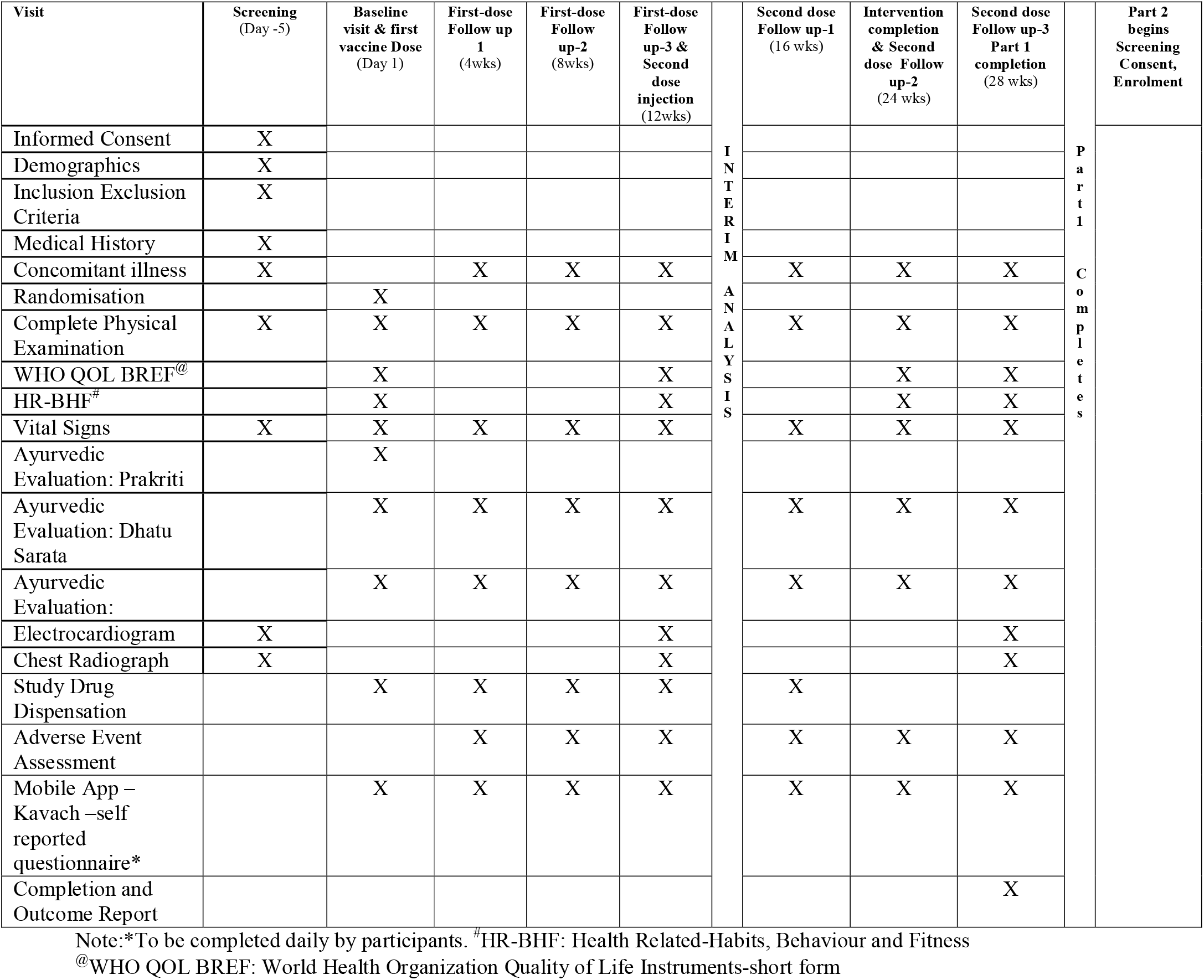
Clinical Assessment Schedule.

**Table 2:** Schedule of Laboratory investigations.

| Visit | Screening<br>(Day -5) | Baseline visit & First vaccine Dose<br>(Day 1) | First dose Follow up 1<br>(4wks) | First dose Follow up-2<br>(8wks) | First-dose Follow up-3 & Second dose injection<br>(12 wks) |  | Second dose Follow up-1<br>(16 wks) | Intervention completion & Second dose Follow up-2<br>(24 wks) | Second dose Follow up-3<br>(28 wks) |  | Part 2 begins Screening Consent, Enrolment |
| --- | --- | --- | --- | --- | --- | --- | --- | --- | --- | --- | --- |
| CBC: Blood Hemoglobin, Total & Differential Count, Platelets Erythrocyte Sedimentation Rate (Westergren) | X |  |  |  | X | INTERIM ANALYSIS |  | X |  | PART 1 COMPLETES |  |
| D-dimer | X |  |  |  | X |  |  | X |  |  |  |
| LFT: Serum Liver Enzymes-SGOT, SGPT, Alkaline Phosphatase Serum Albumin, Globulin, Bilirubin | X |  |  |  | X |  |  | X |  |  |  |
| RFT: Blood Urea Nitrogen Serum Creatinine | X |  |  |  | X |  |  | X |  |  |  |
| Blood Sugar Profile (BSL Fasting & HbA1c) | X |  |  |  | X |  |  | X |  |  |  |
| Urine Routine | X |  |  |  | X |  |  | X |  |  |  |
| +Urine Pregnancy test | X | X | X | X | X |  | X | X | X |  |  |
| #RT- PCR throat, nose swab testing for SARS-CoV2 RNA | X | X | X | X | X |  | X | X | X |  |  |
| SARS- CoV-2 S1, RBD and N Specific IgG |  | X | X | X | X |  | X | X | X |  |  |
| sVNT |  |  |  |  | X |  | X |  | X |  |  |
| SNT (Wuhan & Indian variants) |  |  |  |  |  |  | X |  | X |  |  |

If the vaccinated individual (after first or second dose) contracts COVID 19, then he/she shall be withdrawn from the study and will be treated under the supervision of COVID -19 physician till recovery. The participants who are withdrawn prematurely will be encouraged to continue follow up in a separate group as per the discretion and supervision of the study physician. The latter group will be permitted to use the current protocol methods and analysed separately. All participants will be monitored for safety and immune response and occurrence of any breakthrough SARS-CoV-2 infection. An interim analysis at the end of 12 week is planned to evaluate the effect of Ashwagandha co-administration on safety and immune response following the first dose of the vaccine. The final analysis of data will be carried out after all the participants have completed the study at the end of 28 weeks.

### Recruitment, screening, consent

Apparently healthy volunteers (18-45 years) who will be receiving the COVID-19 vaccine as part of the Government of India vaccination programme will be included in this study. The investigator will be responsible for obtaining written informed consent from each participant prior to screening as per the national ethics guidelines [19]. An information sheet in language best understood by the participant, giving details of the study will be provided to the participant for study and discussion if any with the investigator. Enough time will be given to the participant to make a voluntary decision to participate. Participant will be encouraged to ask questions and clear all doubts in a face to face interview with the investigator. After fully satisfying with the various provisions of the research study the participant will be asked to sign the consent form. Participants will be screened comprehensively by the study physicians for eligibility and to exclude any contraindication to participation in the study. If found eligible, participants will be randomized and enrolled. Participants will be followed up at predetermined time endpoints (8 visits including screening and completion visits) as per protocol till study completion (week 28).

### Eligibility

#### Inclusion criteria

Apparently healthy adults of either sex aged 18-45 years; SARS-COV-2 RT-PCR negative tested volunteers and eligible for vaccination; Written informed consent by the participants; Willing to comply with study protocol requirements; Female participants of childbearing potential with a negative urine pregnancy test carried out within 24 hours prior to study vaccine administration or should be classified as non-childbearing potential (defined as surgically sterile or post-menopausal); Women with a child bearing potential shall continue adequate contraception (Barrier or hormonal) throughout the duration of the study.

#### Exclusion criteria

Any known co-morbidity or acute illness with or without symptoms at the time of study vaccine administration; Participants with a history of COVID -19 any time in the past or currently positive for SARS-CoV-2 by a standard RT-PCR assay; History of known hypersensitivity to any vaccine or any component of the study intervention (Vaccine or Ashwagandha); Any confirmed or suspected condition with impaired/altered function of the immune system or conferring a possible risk of SARS-CoV-2 infection as suggested by investigations (e.g. increased levels of D-dimer)

5. Women who are breast-feeding and pregnant; Individuals who are using any intervention (AYUSH or otherwise) for prophylaxis or with a purpose to increase immunity or influence immune system in any way e.g. steroids, methotrexate, anti-rheumatoid agents, herbal drugs; Individuals who have received systemic immunoglobulins or blood products within 3 months prior to screening; Any contraindication of COVISHIELD™ vaccine

#### Withdrawal criteria

The participants with drug compliance < 70% (estimated by recorded tablet consumption), any serious adverse event, or those with positive RT-PCR tests, or clinical symptoms of COVID-19 will be withdrawn from the study. Withdrawn participants who are RT-PCR positive will be referred to a COVID-19 treatment facility for standard care.

### Randomization

The participants eligible for enrolment will be randomized in a 1:1 ratio to Ashwagandha + COVISHIELD™ or placebo + COVISHIELD™ according to a pre-specified randomization scheme prepared by a statistician (SS) using a computerized system. Block randomization specific to each clinical study site will ensure equal number of participants per group with a small block size (e. g. a block of 4-6 participants). The statistician (SS) will design the randomization schedule following computer generated random numbers. The patient number and the treatment allocation as per the schedule will be printed on a card, which will be sealed in an opaque envelope for each patient. The envelope of each patient will be delivered to respective trial sites. The treatment allocation will be concealed. Considering the study design, it is important to have a dosage form that ensures blinding. The tablets of Ashwagandha and placebo will have identical colour and weight and will be dispensed in sealed bottles. The bottles will have same colour and labelling. The bottles will be specific to the participant. The participant code will be written on each bottle containing the tablets.

#### Monitoring

Following randomization at baseline, participants will be physically checked by study physicians at all visits till the completion of study. At all the time points, assessment of participants will also include in-depth inquiry of possible symptoms of COVID-19 in family members and close associates. The participants will be provided with a user friendly mobile app (named COVID Kavach) designed for COVID-19 studies for daily tracking and monitoring of onset of fever/ respiratory and any other symptoms/vaccine related side effects [20]. The daily response of the participant will be seen by the site physician and co-ordinator and by the overall study co-ordinator; anonymous participant data which will only be identified by a unique study id known to the site investigator will be saved in a dedicated central study server. In addition, a telephonic contact will be made by the site coordinator with the participant once in every two weeks till study completion to find out about any matter concerning health or the study. The schedule of enrolment, interventions, and assessments is provided in **Table 3**.

**Table 3:**
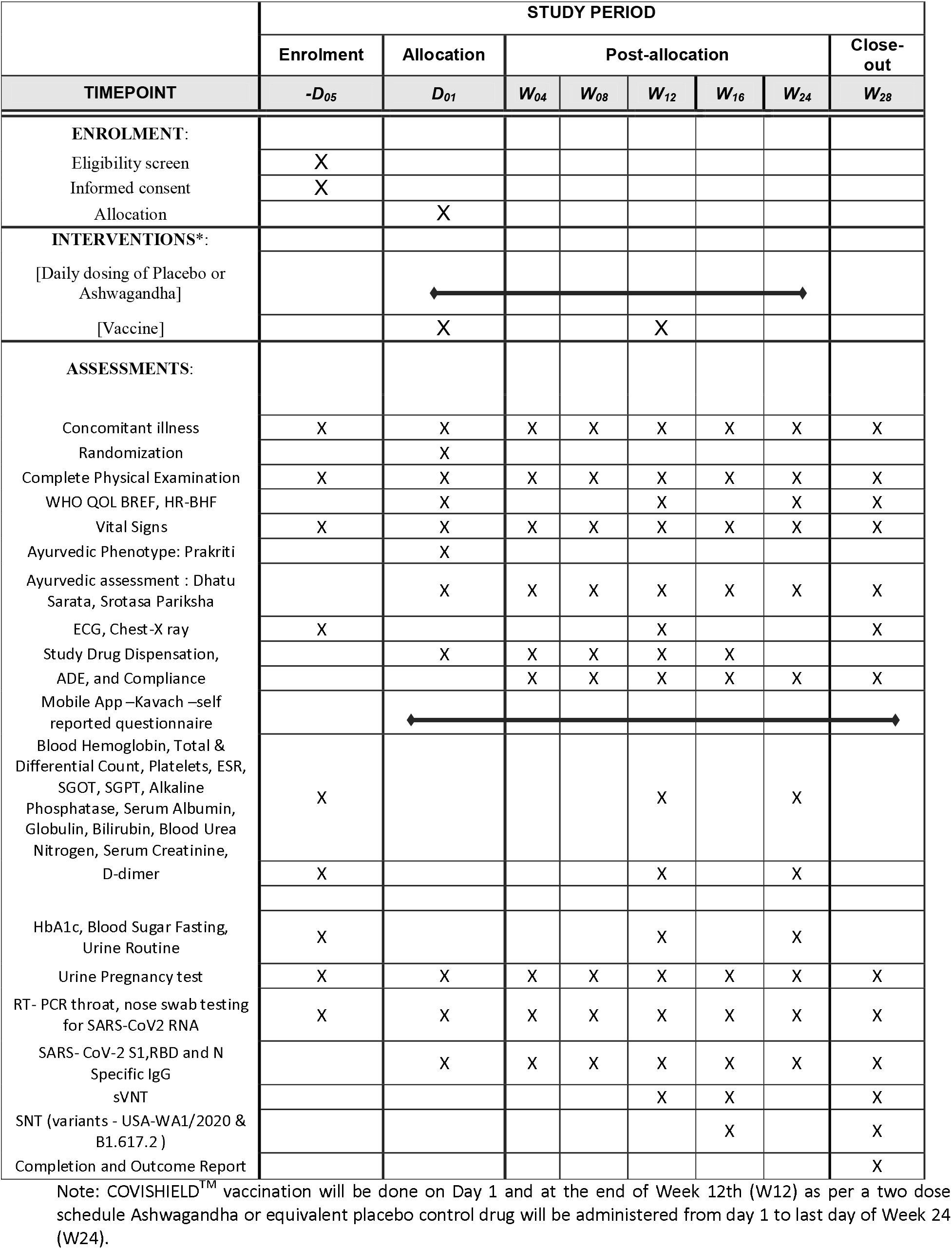
Schedule of enrolment, interventions, and assessments.

#### Compliance

The participants will be daily monitored for fever, respiratory and any other symptoms and vaccine related side effects. Telephonic follow up will be done once in every two weeks throughout the study. The study coordinator will contact the participants for monitoring symptoms and ensure compliance. Medication logs will be maintained for both vaccine use and Ashwagandha tablet consumption. On each return visit, the participant will be instructed to bring all unused medication and a tablet count will be performed and recorded in the medication log. COVID-Kavach App and regular telephonic contact will be used to remind participants to adhere to study medication instructions.

### Trial Intervention

The choice of competitors includes two arms as Ashwagandha and Placebo in addition to vaccine. The participants from placebo arm will have immunogenic effects only of vaccine, whereas participants in Ashwagandha + Vaccine arm may have additional safety, immunogenicity and protection from disease. Thus, the placebo arm will serve as a control to ascertain adjuvant effects offered by Ashwagandha.

Arm 1: Ashwagandha tablets + Vaccine (n=600): COVID-19 vaccine (COVISHIELD™) will be administered as per the 2-dose schedule on Day 1 and at the end of 12 weeks as 0.5 ml dose intramuscularly as per the national guideline. Dose of Ashwagandha prescribed for this study is one tablet twice daily administered from 1st vaccine dose (Day 1) and continued up to 24 weeks. Each tablet used in this trial shall contain aqueous extract equivalent to 4 grams Ashwagandha root powder.

Arm 2: Placebo tablets + Vaccine (n=600): COVID-19 vaccine (COVISHIELD™) will be administered as per the 2-dose schedule on Day 1 and at the end of 12 weeks as 0.5 ml dose intramuscularly as per national guideline. Twice daily dose of placebo tablets will be administered from 1st vaccine dose and continued up to 24 weeks. The appearance and weight of placebo tablet (containing microcrystalline cellulose) will be same as that of Ashwagandha tablet.

The COVID-19 vaccine will be administered in a two-dose schedule (0.5ml per dose) intramuscularly as per standard practise in India. One tablet of Ashwagandha or placebo will be administered twice daily (on empty stomach in the morning and after dinner) with water. Quality of Ashwagandha roots is ensured in accordance with prescribed pharmacopoeia standards [21] A window period of 7 days will be allowed on either side of the scheduled visit for exceptional or any other anticipated reasons at the discretion of the investigator. The window period for second dose of vaccine will be two weeks.

#### Clinical outcomes

**Primary Outcome Measure:** Immunogenicity as measured by SARS-CoV-2 spike (S1) and RBD-specific IgG antibody titres. **Secondary Outcome Measures:** Safety as measured by occurrence of adverse events. Protective immune response as measured by SARS-CoV-2 specific neutralising antibody titer; Duration of sustenance of SARS-CoV-2 virus neutralizing antibody titre; percent seroconversion after each vaccine dose; number of participants with SARS-CoV-2 breakthrough infections; proportion of participants with COVID 19 (RT PCR assay confirmed) after the first dose and both showing evidence of infection before vaccination (Time frame: Day 1 vaccination to Week 12 or second dose and whichever is earlier); proportion of participants with COVID-19 (RT-PCR assay confirmed) after completion of the second dose (Time frame: Second dose to week 28 or study completion) and Quality of life measured using standard WHO Quality Of Life-BREF 1997 [22] and recently designed and validated Health Related - Behaviour Habit Fitness (CRD Pune version 2020) questionnaires. All clinical and laboratory investigations and assessments pertaining to general health, and to rule out concurrent medical illness including COVID-19 will be carried out as per **Table1** and **Table 2**.

#### Adverse events

All adverse events (AEs) and adverse events following immunization (AEFI) occurring during the study will be recorded and monitored as per ICH-GCP (2016) [23] and ICMR guidelines [19]. Safety would also be assessed in case of all withdrawals. All adverse events will be recorded and meticulously followed till resolution. Serious adverse events will require immediate reporting by the study investigator to the Sponsor and Ethics Committee, within 24 hours, of becoming aware of the SAE. All Adverse Events will be classified by the study physician and recorded in the AE form. Any symptoms recorded on COVID-Kavach App will be immediately followed by investigators to rule out COVID-19. The Data Safety Monitoring Board (DSMB) will be formed to monitor safety of the participants and assess safety data. Clinical assessments and lab investigations will be carried out as per **Table 1** and **Table 2**.

#### Concomitant Medication

As a general principle, concomitant medication of any kind will be kept to a minimum and will necessarily require clearance from a primary care physician and with full information provided to the study physician. All concomitant medication will be recorded in the study case record form (CRF) by the study physician. All complete CRFs/eCRF duly signed by the investigator and counter signed by principal investigator will be submitted within two weeks of the completion of the study to the Chief Clinical Coordinator. Although the study includes healthy participants, the medication required for any new illness during the study will be taken under supervision of the primary care physician and meticulously recorded in the participant CRF. Participants will be permitted general symptomatic medications like paracetamol and non-steroidal anti-inflammatory drugs for short period (<4 days) and including for any untoward mild to moderate symptoms like headache and body aches following vaccination. Patients will be carefully explained not to consume any other herbal and home remedies. Patients will be advised not to use any kind of food and nutrition supplement. Similarly, participants will be advised not to consume any kind of vitamins and/or minerals without a specific advice of physician.

### Statistical analysis

#### Sample size and Power

The sample size of 1200 (600 in each group) participants is calculated based on the primary objective of improved immunogenicity of the vaccine with coadministration of Ashwagandha. Also, it was felt important to obtain a reasonably suitable (non-probabilistic) sample size for safety and tolerability. As there was no prior data on the expected effect size of the difference between the two study groups, a superiority margin of 7% was decided by expert consensus between the biostatistician (SS) and senior authors (AC, GT, PC and BP). The sample size was the calculated with 80% power and after consideration of interim analysis at the end of 12 weeks; assuming vaccine efficacy of 80% [24]. The calculated sample size with the provision of one interim analysis was rounded to 600 subjects per each of the two study arms and totalling 1200.

#### Analysis

A standard statistical software package will be used for statistical tests (parametric and non-parametric). The analysis will be both intention-to-treat (ITT) and as per-protocol (PP). All randomized participants will be included in the safety analysis. All statistical processing will be performed using standard statistical package (SPSS, BMDP, and CIA) unless otherwise stated. The data will be individually analysed for central tendencies (mean, median), range, standard deviation and 95% confidence intervals for each intervention arm in the study. Titre data will be ‘log’ transformed before any analyses [and GMT, GSD, etc. will be reported & will be used for testing with respect to that parameter. Data will be tabulated and graphically shown using standard format using MS Excel and other software programs. Standard parametric (Student t-tests – paired and unpaired, ANOVA, etc.) and non-parametric statistical tests (Wilcoxon Signed Rank test, Mann-Whitney ‘U’ test, Kruskal-Wallis test, etc.) will be used depending upon the normality of the data, level of measurement and appropriateness/applicability. Fisher’s exact test or Chi square will be used to test the proportion of participants classified as efficacy failures and those with AE. All AEs will be presented as proportion and with Wilson’s recommended exact binomial 95% Confidence Interval.

#### Electronic database

An e-database will be created to capture all the important variables required for the final study report and supervised by the CRO. Access to e-database will be limited to the site investigator (only for the particular site), study co-ordinators and project team and CRO, and will be password protected. The CRO will be responsible for the security and back up of the study e-database. The database will be checked for missing data and inconsistencies by the CRO from time to time in close liaison with the site investigator. The CRO will lock the database after informing the clinical coordinator and sponsor and then submit copy for statistical analysis and report writing both for interim and final report.

#### Trial care and record retention

The participants will be advised to continue the usual protective measures like physical distancing, respiratory and hand hygiene, and wearing masks. They will be advised to follow guidelines of Ministry of AYUSH for COVID-19 prophylaxis. All clinical study documents will be retained by the investigator for at least 5 years after the study and await instructions of their final disposal from the sponsor. All protocol amendment will be approved by the respective Ethics Committees before implementation. In the event of an emergency of any type, medical or otherwise, the site investigator will inform the appropriate Ethics Committee, CRO, study coordinators and sponsor within 24 hours. The Sponsor (CCRAS) will provide insurance coverage with respect to any liability caused by the investigational products and or study related procedures in connection with the clinical study. The terms and conditions will apply as specified in the insurance policy document. The quantum of compensation for the Serious Adverse Events leading to the death will be given to the study participant/patients’ nominee as per recommendations of the EC, DSMB, Central Drugs Standard Control Organisation (CDSCO) expert committee and as per the regulatory guidelines.

## DISCUSSION

Several vaccines have been approved in the first year of the pandemic on priority for an emergency use. We still do not have a clinically acceptable specific drug to treat COVID-19. The case fatality rate in India is still hovers around 2% in the second pandemic year. It is indeed challenging to predict disease progression at an early stage. Agreeably, vaccination is reassuring, however the future remains somewhat uncertain. The success with vaccines is influenced by several host and environmental factors. The data on safety, tolerance and efficacy is not adequate. In case of natural COVID-19 infections it has been observed that the titers of IgG against spike protein may last for about 6 months, while the immunological memory may lasts up to 8 months [25] [26]. However, the duration of protection after vaccination has not been established so far. A looming fear of more waves may sweep away some of the benefits of the vaccination in the population. The world indeed is immensely stressed and mental health is at stake [27]. How to improve the performance of the vaccines remains a pivotal question. The current study protocol is planned and designed against this perspective.

Previous clinical studies have highlighted the role of humoral as well as cell-mediated immune responses in recovery from SARS-CoV-2 infection [28] [29]. In previous studies the selective-Th-1 upregulating activity of Ashwagandha was demonstrated and in infection models, co-administration of Ashwagandha with DPT vaccine resulted in improved antibody titres against lethal challenge of pertussis toxins [10]. *In silico* network ethnopharmacological investigations also suggest the ability of bioactive compounds of Ashwagandha to modulate immune pathways involved in innate and adaptive immune responses [30] [31]. Further, the interim efficacy data for ChAdOx1 nCoV-19, evaluated in four trials across UK, Brazil and South Africa shows a protection of 64·1% after one standard dose [32]. COVISHIELD™ has been administered in two doses to a large population in India. We hypothesise that co-administration of Ashwagandha with the COVISHIELD™ vaccine may enhance the antibody titres particularly after the first dose of the vaccine. A sustained long-term immune response to the vaccine has implications for prevention of reinfection. Therefore, the effect of Ashwagandha on the sustenance of neutralizing antibody titres up to four months after the second dose will also be evaluated in this study [33]. Ashwagandha is also known to improve overall mental wellbeing.

Our trial has the potential to have a significant impact on current vaccination program by improving immunogenicity, protection and general wellbeing with simple, safe and affordable intervention. In this trial we expect a beneficial effect beginning soon after the first dose of vaccination. We hope to investigate biological plausibility that the immune protection offered by a long-term co-administration of Ashwagandha may effectively address some of the vexing issues associated with the vaccination.

## Data Availability

The status of the study and the results (after final analysis) will be uploaded on CTRI Website URL - http://ctri.nic.in.

http://www.ccras.nic.in/

## ETHICS AND DISSEMINATION

This trial protocol is designed in accordance SPIRIT guidelines [34]. This trial will be conducted and reported in accordance with the International Conference on Harmonization Harmonized Tripartite Guidelines for Good Clinical Practice, the Declaration of Helsinki, CONSORT guidelines and applicable government regulations. This protocol has been duly registered with Clinical Trial Registry – India (CTRI), Indian Council of Medical Research – National Institute of Medical Statistics, New Delhi. Registration Number: CTRI/2021/06/034496. Date of Registration June 30, 2021 - Trial Registered Prospectively. The status of the study and the results (after final analysis) will be uploaded on CTRI Website URL - http://ctri.nic.in. The study results will be published in scientific journals, also be disseminated to the public through electronic, social and print media.

## Acknowledgment

A special thanks to Vaidya Dr Rajesh Kotecha, Secretary, Ministry of AYUSH, Government of India, for his invaluable guidance and inspiration. We thank Vaidya KS Dhiman, former DG and other non-author contributors from the CCRAS project team - Ravindra Singh, Shruti Khanduri, Sophia Jameela, Amit Kumar Rai, Azeem Ahmed, B S Sharma. We also thank all reviewers who helped in improving the quality of the protocol.

## References

1 Vabret N, Britton GJ, Gruber C, et al. Immunology of COVID-19: Current State of the Science. Immunity 2020;52:910–41. doi:10.1016/j.immuni.2020.05.002

2 Knoll MD, Wonodi C. Oxford-AstraZeneca COVID-19 vaccine efficacy. Lancet (London, England) 2021;397:72–4. doi:10.1016/S0140-6736(20)32623-4

3 Poonia B, Kottilil S. Immune Correlates of COVID-19 Control. Front Immunol2020;11:569611. doi:10.3389/fimmu.2020.569611

4 Ewer KJ, Barrett JR, Belij-Rammerstorfer S, et al. T cell and antibody responses induced by a single dose of ChAdOx1 nCoV-19 (AZD1222) vaccine in a phase 1/2 clinical trial. Nat Med 2021;27:270–8. doi:10.1038/s41591-020-01194-5

5 Srikanth N, Rana R, Singhal R, et al. Mobile App-Reported Use of Traditional Medicine for Maintenance of Health in India During the COVID-19 Pandemic: Cross-sectional Questionnaire Study. JMIRx med 2021;2:e25703. doi:10.2196/25703

6 Sharma R, Dash B. Charaka Samhita. Choukhamba Sanskrit Series Office, Varanasi 2000.

7 Tillu G, Salvi S, Patwardhan B. AYUSH for COVID-19 management. J Ayurveda Integr Med 2020;11:95–6. doi:10.1016/j.jaim.2020.06.012

8 Patwardhan B, Chavan-gautam P, Gautam M, et al. Ayurveda rasayana in prophylaxis of COVID-19. Curr Sci 2020;19:1158–60. doi:https://www.currentscience.ac.in/Volumes/118/08/1158.pdf

9 Bani S, Gautam M, Sheikh FA, et al. Selective Th1 up-regulating activity of Withania somnifera aqueous extract in an experimental system using flow cytometry. J Ethnopharmacol 2006;107:107–15. doi:10.1016/j.jep.2006.02.016

10 Gautam M, Diwanay SS, Gairola S, et al. Immune response modulation to DPT vaccine by aqueous extract of Withania somnifera in experimental system. Int Immunopharmacol 2004;4:841–9. doi:10.1016/j.intimp.2004.03.005

11 Raut AA, Rege NN, Tadvi FM, et al. Exploratory study to evaluate tolerability, safety, and activity of Ashwagandha (Withania somnifera) in healthy volunteers. J Ayurveda Integr Med 2012;3:111–4. doi:10.4103/0975-9476.100168

12 Verma N, Gupta SK, Tiwari S, et al. Safety of Ashwagandha Root Extract: A Randomized, Placebo-Controlled, study in Healthy Volunteers. Complement Ther Med 2021;57:102642. doi:10.1016/j.ctim.2020.102642

13 Agnihotri AP, Sontakke SD, Thawani VR, et al. Effects of Withania somnifera in patients of schizophrenia: a randomized, double blind, placebo controlled pilot trial study. Indian J. Pharmacol. 2013;45:417–8. doi:10.4103/0253-7613.115012

14 Patel SB, Rao NJ, Hingorani LL. Safety assessment of Withania somnifera extract standardized for Withaferin A: Acute and sub-acute toxicity study. J Ayurveda Integr Med 2016;7:30–7. doi:10.1016/j.jaim.2015.08.001

15 Pires N, Gota V, Gulia A, et al. Safety and pharmacokinetics of Withaferin-A in advanced stage high grade osteosarcoma: A phase I trial. J Ayurveda Integr Med 2020;11:68–72. doi:10.1016/j.jaim.2018.12.008

16 Saggam A, Limgaokar K, Borse S, et al. Withania somnifera (L.) Dunal: Opportunity for Clinical Repurposing in COVID-19 Management. Front Pharmacol 2021;12:623795. doi:10.3389/fphar.2021.623795

17 Folegatti PM, Ewer KJ, Aley PK, et al. Safety and immunogenicity of the ChAdOx1 nCoV-19 vaccine against SARS-CoV-2: a preliminary report of a phase 1/2, single-blind, randomised controlled trial. Lancet (London, England) 2020;396:467–78. doi:10.1016/S0140-6736(20)31604-4

18 Ramasamy MN, Minassian AM, Ewer KJ, et al. Safety and immunogenicity of ChAdOx1 nCoV-19 vaccine administered in a prime-boost regimen in young and old adults (COV002): a single-blind, randomised, controlled, phase 2/3 trial. Lancet (London, England) 2021;396:1979–93. doi:10.1016/S0140-6736(20)32466-1

19 National guidelines of Ethics Committees reviewing biomedical and health research during COVID-19 pandemic. 2020. https://main.icmr.nic.in/sites/default/files/guidelines/EC_Guidance_COVID19_06_05_2020.pdf

20 Covid Kavach -Apps on Google Play. https://play.google.com/store/apps/details?id=com.moa.covidrecorder&hl=en (accessed 26 Jun 2021).

21 Ayush Dept, MHFW G. The Ayurvedic Pharmacopoeia of India Part-I Volume-I. 1989. http://www.ayurveda.hu/api/API-Vol-1.pdf (accessed 5 Feb 2020).

22 WHOQOL-BREF Introduction, Administration, Scoring and Generic Version of The Assessment Field Trial Version December 1996 Programme on Mental Health. Published Online First: 1996.https://www.who.int/mental_health/media/en/76.pdf

23 ICH GCP - ICH harmonised guideline integrated addendum to ICH E6(R1): Guideline for Good Clinical Practice ICH E6(R2) ICH Consensus Guideline. https://ichgcp.net/ (accessed 1 Jul 2021).

24 Sarmukaddam S. Biostatistical Methods for Medical Research. 2019. https://rfppl.co.in/bookdetailsinformation.php?mid=2&id=74

25 Dan JM, Mateus J, Kato Y, et al. Immunological memory to SARS-CoV-2 assessed for up to 8 months after infection. Science 2021;371. doi:10.1126/science.abf4063

26 Ma H, Zeng W, He H, et al. Serum IgA, IgM, and IgG responses in COVID-19. Cell Mol Immunol 2020;17:773–5. doi:10.1038/s41423-020-0474-z

27 Xiong J, Lipsitz O, Nasri F, et al. Impact of COVID-19 pandemic on mental health in the general population: A systematic review. J Affect Disord 2020;277:55–64. doi:10.1016/j.jad.2020.08.001

28 Chen G, Wu D, Guo W, et al. Clinical and immunological features of severe and moderate coronavirus disease 2019. J Clin Invest 2020;130:2620–9. doi:10.1172/JCI137244

29 Gudbjartsson DF, Norddahl GL, Melsted P, et al. Humoral Immune Response to SARS-CoV-2 in Iceland. N Engl J Med 2020;383:1724–34. doi:10.1056/NEJMoa2026116

30 Chandran U, Patwardhan B. Network ethnopharmacological evaluation of the immunomodulatory activity of Withania somnifera. J Ethnopharmacol 2016;197:250–6. doi:10.1016/j.jep.2016.07.080

31 Borse S, Joshi M, Saggam A, et al. Ayurveda botanicals in COVID-19 management: An in silico multi-target approach. 2021. doi:10.1371/journal.pone.0248479

32 Voysey M, Costa Clemens SA, Madhi SA, et al. Single-dose administration and the influence of the timing of the booster dose on immunogenicity and efficacy of ChAdOx1 nCoV-19 (AZD1222) vaccine: a pooled analysis of four randomised trials. Lancet (London, England) 2021;397:881–91. doi:10.1016/S0140-6736(21)00432-3

33 Muruato AE, Fontes-Garfias CR, Ren P, et al. A high-throughput neutralizing antibody assay for COVID-19 diagnosis and vaccine evaluation. Nat Commun 2020;11:4059. doi:10.1038/s41467-020-17892-0

34 Chan A-W, Tetzlaff JM, Gøtzsche PC, et al. SPIRIT 2013 explanation and elaboration: guidance for protocols of clinical trials. BMJ 2013;346:e7586. doi:10.1136/bmj.e7586

